# Secondary causes among adult patients presenting with first-episode psychosis to acute medical settings in Hong Kong: A 10-Year retrospective study

**DOI:** 10.64898/2026.09.02.26362044

**Authors:** Steven Wai Ho Chau, Charis KW Lam, Matthew PM Yu, Yuen Cheuk Wong, Joseph CH Choi, Howan HW Leung

## Abstract

**Background:** The prevalence and pattern of secondary causes of first-onset psychosis (FEP) in Asian populations are understudied. Our objectives are to investigate the prevalence and pattern of secondary causes of FEP presenting in an acute medical setting in a metropolitan, Chinese-predominant population, and to investigate the prevalence and pattern of undiagnosed conditions presenting as psychosis.

**Method:** This is a retrospective observational study. We reviewed medical records of patients referred to the consultation psychiatry team at a tertiary acute teaching hospital in Hong Kong from January 2015 to Apr 2025. The inclusion criteria of the study are 1) age 18-64 at the time of the assessment, and 2) FEP confirmed by the consultation liaison team. Patients diagnosed with delirium were excluded.

**Result:** Among the 384 patients included in the study (mean age = 40.0 ±13.7 years, 69% female), secondary causes of psychosis were found in 9.6% (n=37) of the cohort. Substance use is the most common secondary cause found (n=11, 2.9% of all FEPs) overall. Among those with secondary psychoses, patients’ underlying conditions were revealed only by the workup in relation to the FEP in 16 of them, of which definitive or probable autoimmune encephalitis (n=5) and early-onset dementia (n=4) were the most common conditions uncovered.

**Conclusion:** The prevalence of secondary psychoses in our FEP cohort is lower than published international figures. Clinicians need to be aware of the suspicious clinical features suggestive of autoimmune encephalitis and early-onset dementia in FEP patients with otherwise unremarkable past history and toxicology test.

## Background

First-episode psychosis (FEP) is a critical diagnostic and management window. Medical practitioners, including psychiatrists and non-psychiatrists, are often reminded to consider underlying secondary causes (or so-called “organic” causes) in any FEP, because the workup and treatment pathways for those with such causes differ substantially. In some cases, timely discovery of undiagnosed occult causes, e.g. autoimmune encephalitis or intracranial infection, is prognosis-determining, making the diagnosis a matter of medical urgency. A recent meta-analysis estimated that about 14% of FEPs have secondary causes, with substance abuse being by far the most common cause(1). However, several important study limitations require cautious interpretation of the results. First, the studies included in the meta-analysis were mostly conducted in Western countries, with only one study from Asia (Japan)(2). In addition, that particular Japanese study only looked at psychosis caused by autoimmune encephalitis, without looking into other medical causes. Therefore, there are no data on the prevalence and distribution of secondary causes of FEP in Asian populations. Variation in behavioural (e.g. prevalence of substance abuse) and disease patterns(e.g. prevalence of autoimmune disease, infection) may create global variation in patterns in secondary causes of FEP, which is evidenced by Blackman et al’s analysis that showed that African countries had a higher prevalence of medical causes among psychotic patients compared to the rest of the cohort. Second, we have no data on the prevalence of previously undiagnosed or undocumented medical conditions presenting as psychosis, which is arguably an even more important clinical subcategory.

The primary objective of the current study is to investigate the prevalence and pattern of secondary causes of FEP presenting in an acute medical setting in a metropolitan, Chinese-predominant population. The secondary objective is to investigate the prevalence and pattern of undiagnosed conditions presenting as psychosis.

## Method

This is a retrospective observational study. We reviewed medical records of patients referred to the consultation psychiatry team at a tertiary acute teaching hospital in Hong Kong, serving a catchment population of about 700,000, from January 2015 to Apr 2025. The consultation liaison psychiatric team received referrals from the Accident and Emergency Department and other inpatient units. The inclusion criteria of the study are 1) age 18-64 at the time of the assessment, and 2) FEP, defined as the first lifetime psychotic episode regardless of aetiology, was confirmed by the consultation liaison team. Patients diagnosed of delirium were excluded. We reviewed the patient’s medical record at the time of assessment, and their subsequent treatment and investigation progress, if available. A senior psychiatrist determined the final diagnosis and potential underlying medical cause after considering the treating team’s original diagnosis and the patient’s subsequent clinical course.

## Results

We screened 426 consecutive referrals, and 384 were included in the study (mean age = 40.0 ±13.7 years, 69% female, 97% ethnic Chinese). Almost all patients received blood tests and brain imaging (CT or MRI brain), 319 had documented urine toxicology tests, and 103 received lumbar puncture. Overall, underlying medical causes that can explain the onset of psychosis was found in 9.6% (n=37) of the sample: 11 were substance-related (2.9%), 6 were explained by neurodegenerative diseases (1.3%), 3 had definitive autoimmune encephalitis (0.8%; 2 anti-NMDA receptor type and 1 anti-LGI-1 type), 2 had probable autoimmune encephalitis which responded to immunotherapy(0.5%) (according to criteria by Graus et al. (3)), 5 had underlying neurodevelopmental abnormalities or epilepsy (1.3%), 3 were related medication side effect (0.8%), 2 were presumed to be COVID-related (0.5%), and 5 had other diagnoses. (The brief descriptions of these cases can be found in the Supplementary material). Among them, their secondary causes were documented in their past medical history prior to the onset of psychosis in 21 patients, while 16 patients’ underlying conditions were revealed only by the workup in relation to the FEP (5 were diagnosed with definitive or probable autoimmune encephalitis after workup, 4 were diagnosed with neurodegenerative diseases after workup or subsequent observation of clinical course (early-onset dementia of a mix of aetiologies), 3 had previously undocumented substance use that was uncovered by toxicology tests, 1 was found to have COVID upon psychosis presentation, and 1 was found to have intracranial mass by brain imaging). Among the 5 patients with definitive and probable or definitive autoimmune encephalitis (age range: 29-59 years, 4 females), 4 had sudden onset of psychosis (< 2 weeks), 4 had cognitive impairment upon presentation, and 4 had motor abnormalities or seizures at some point in their course of illness. Among the 4 patients with early-onset dementia presenting as FEP (age range: 54-60 years, 4 males), all complained of cognitive decline at the time of presentation.

## Discussion

To the best of our knowledge, this is the first study on the secondary causes in patients presenting with FEP to acute medical settings in a predominantly Chinese population. The overall frequency was lower than the prevalence reported by a meta-analysis by Blackman et al (9.6% vs 14%).

In line with the international trend, substance use is the most common cause of secondary psychosis in our cohort. However, the prevalence of substance use in our cohort is very low compared to Blackman et al’s data (13% among all psychoses; no FEP-specific data, however)(1). It is consistent with the lower prevalence of substance use disorder in East Asia, compared to the rest of the world (4). Rare cases of psychoses associated with COVID were observed in our cohort. While COVID-associated psychosis is widely observed, the diagnostic criteria and its clinical course are not well established. Therefore, the diagnoses of COVID-associated psychosis we made are tentative and based on the temporal sequence of events. It is noted that the majority (57%) of secondary causes in the cases were documented before the presentation of psychosis, meaning that a careful history taking of past medical history or substance use would suggest the causes in most of the cases. However, patients with autoimmune encephalitis or early-onset dementia more commonly present with a ‘clean’ history, which requires a clinician’s high index of suspicion and recognition of the typical demographics and course of illness of these differential diagnoses to triage appropriate patients for further workup, such as cerebrospinal fluid autoantibody tests or functional brain imaging. Our study did not systematically collect detailed psychopathology, as limited by retrospective data collection from available clinical notes. While Blackman et al (2026) suggested that psychopathology, such as visual hallucinations and poverty of speech, is more common in secondary psychoses and have clinical value in screening out these cases(5), our data suggest that recognition of patients’ medical (including co-presenting neurological and cognitive symptoms) and substance use history, neurological examination, and urine toxicology tests can effectively screen out the majority of those at risk of secondary psychosis for further workup in an adult setting. The key strength of our study is a representative sample obtained through consecutive sampling that includes all possible cases. Readers can also refer to our supplementary material to verify individual cases’ diagnoses. Limitations include a modest sample size, an arbitrary age cut-off due to the scope of service provision, and referral bias. In many cases, we cannot definitively establish cause-and-effect relationships.

## Conclusion

The prevalence of secondary causes of FEP in our Chinese-predominant cohort from an acute medical setting in Hong Kong is lower than published international figures, mostly because of a lower prevalence of substance-related psychosis. Most secondary causes of FEP were known from patients’ history. Definitive or probable autoimmune encephalitis and early-onset dementia are the most common secondary causes that present as psychosis at their first medical contact.

## Data Availability

All data produced in the present study are available upon reasonable request to the authors.

## Patient consent for publication

Not applicable.

## Ethics approval

The Joint CUHK-NTEC Clinical Research Ethics Committee approved the study (ref: 2024.606).

## Acknowledgment

We thank Ms Yee Lok Lai and Ms Tsz Ching Lam for assisting us in data collection.

