## Supplementary material for "Secondary causes among adult patients presenting with first-episode psychosis to acute medical settings in Hong Kong: A 10-Year retrospective study": Supplmentary material

| Case no. | Daignosis | Brief clinical description | Psychosis as presenting symptoms of underlying physical cause? |
|---|---|---|---|
| 1 | Alcohol induced psychotic disorder | M/History of chronic alcohol abuse | N |
| 2 | Chemotherapy induced psychosis | M/ presented with 2 weeks of acute onset presecutory delusion and aggressiveness, two weeks after change of targeted therapy for lung cancer (Lorlatinib). Symptoms resolved in two weeks after stopping Lortatinib without need for antipsychotics. No recurrance in the next 2 years. | N |
| 3 | Definite autoimmune encephalitis (anti NMDAR) | M/ Presented with an acute onset of cognitive impairment, agitation, and insomnia for 2 weeks, followed by the onset of auditory hallucination 2 days before hospitalisation. CSF analysis showed marked pleocytosis without evidence of infection. MRI brain showed abnormal T2W signal over left temporal lobe, suggestive of encephalitis. Anti-NMDAr +ve in CSF | Y |
| 4 | Definite autoimmune encephalitis (anti-LGI-1) | F/Presented with VH and significant congitive impairment for 1 month. Subtle seizure episode was noted later captured by video EEG (episodes of motor arrest and impaired awareness, panic/depersonalization feeling) . MRI with contrast suggestive of encephalitis. Confirmed anti LGI-1 autoimmune encephalitis by CSF autoantibodies testing 3 months after presentation. Treated by high dose pulse steroid. | Y |
| 5 | Definite autoimmune encephalitis (anti-NMDAR) | F/presented with abrupt-onset insomnia, cognitive impairment, and auditory and visual hallucinations within a week. She subsequently developed orofacial dyskinesia and seizures in the following two weeks. While the MRI of the brain with contrast did not reveal encephalitic changes, the CSF analysis showed marked pleocytosis without evidence of infection. Anti-NMDAr +ve in CSF | Y |
| 6 | Early-onset dementia | F/presented with subacute cognitive decline and VH. CT brian: bil parietal atrophy. SPECT: hypoperfusion with AD pattern | Y |
| 7 | doperminergic drug induced psychosis | M/with known Parkinson's disease on dopaminergic medication. He presented with 1 week hx of VH and agitation, which resolved after witholding PD med | N |
| 8 | Early-onset dementia | F/ presented with few years of gradual cognitive decline abd 9 months of insidious onset of persecuotry and infidelity delusion. MRI: progressive cortical and right hippocampal atrophy | Y |
| 9 | Early-onset dementia | F/ she was admitted for months of cognitive decline and self neglect first, and mentioned AH later. She was initially diagnosed late onset schizophrenia but subseqeuntly she was noted to have progressive cognitie and motor deline. SPECT: highly supicious of DLB pattern with mixed vascular dementia | Y |
| 10 | Early-onset dementia | F/ with history of anxiety disorder, presented with 3 weeks of rapid cognitive decline, followed by auditory hallucination and persecutory delusion. Subsequently develop further cognitive decline and Parkinsonism feature. SPECT scan suggested DLB-like hypoperfusion pattern. | Y |
| 11 | Parkinson's disease | M/ Known advanced Parkinson's disease, presented with months of delusion of infestiation. Resolved after reducing dopaminergic drug and low dose quetiapine (25mg). | N |
| 12 | Post-ictal psychosis | F/ with known refractory epilepy since childhood. Report 2 days of AH and paranoid idea after a breakthrough GTC. No antipsychotics prescribed. No subseqeunce recurrence of psychosis/ | N |
| 13 | Post-ictal psychosis/neurodevelopmental | M/ Known left schizencephaly with epilepsy. Presented with new onet delusion of infestation after breakthrough seizure. | N |

| | | | |
|---|---|---|---|
| 14 | Probable autoimmune encephalitis | F/Probable AE as : 1 month onset, presence of psychosis, seizure, abnormal movement, CSF pleocytosis, infection excluded. Full recovery. extended AB panel -ve. Malignancy -ve. | Y |
| 15 | Probable autoimmune encephalitis | F/Probable AE as 1 week onset of psychotic symptoms, confusion, autonomic instability, abnormal movement.CSF analysis showed WCC up to 197, but no autoimmune antibody was detected. Psychotic symptoms did not respond to antipsychotics, but resolved with aggressive immunotherapy. | Y |
| 16 | Psychosis related to vitamin B12 deficiency | M/ presented with 5 years of hypochrondrical delusion. Found to have very low vitamin b12 level upon presentation. Long term B12 replacement was prescribed. Antipsychotic-free subsequently. | Y |
| 17 | Psychosis with underlying congenital brain malformation | M/multiple congential brain malformation including septo-optic dysplasia and partial callosal agenesis. Epilepsy, Present ed with 7 years of low grade AH and VH which did not require antipsychotic treatments. | N |
| 18 | Psychosis with underlying epilepsy, oral-facial-digital syndrome | M/with known oral faical digital syndrome, epilepsy and learning disability. He presented with 2-3 months hx of persecutory delusion and subsequent psychomotor disturbance | N |
| 19 | Psychosis with underlying multiple sclerosis | F/ with known multiple sclerosis. Presented with 5 years of AH, persecuotry delusion and progressive cognitive decline and gait instability. | N |
| 20 | Psychosis with underlying tuberous sclerosis | F/ with known tuberous sclerosis. She presented with 4 years of auditory hallucination and referential delusion and severe suicidal attempt. | N |
| 21 | Steroid induced manic psychosis | F/ with underlying SLE nephritis. She presented with acute manic psychosis within days of doubling the dose of prednisolone, which resolved within a weekafter the steroid dose was reduced. No antipsychotics used. | N |
| 22 | Substance induced psychosis | n/a | N |
| 23 | substance induced psychosis (Cocaine) | n/a | N |
| 24 | substance induced psychosis (Cocaine) | chronic cocaine use | N |
| 25 | substance induced psychosis (methamphetamine) | Known chronic methamphetamine use | N |
| 26 | substance induced psychosis (methamphetamine) | Initially diagnosed of psychotic disorder with no known substance abuse history. Urine toxicology revealed methamphetamine use. | Y |
| 27 | substance induced psychosis (sibutramine) | F/ Acute psychosis. urine toxicology: sibutramine. Patient's use of sliming pills were not known before, nor did the patient knew about the ingredients | Y |
| 28 | substance induced psychosis (sibutramine) | No documented hx of sliming pill use - but later revealed by urine toxicology result | Y |
| 29 | substance induced psychosis (THC) | THC in urine, Known case of substance disorder clinic | N |
| 30 | Substance induced psychosis (THC) | THC in urine, Known case of substance use disorder | N |
| 31 | Substance induced psychosis (THC) | THC in urine, Known case of substance use disorder | N |
| 32 | suspected COVID-related psychosis | M/ Patient was diagnosed with COVID, followed by abrupt onset of psychosis 2 days later. His psychosis was treatment-resistant initially, requiring clozapine use. However the psychosis then resolved in 6 months despite clozapine was changed to relatively low dosage of atypical antipsychotics (limited by side effect). He had another psychotic relapse upon another respiratory infection. No family hx of psychosis. | N |
| 33 | suspected COVID-related psychosis | F/hx of mild depression but drug free for long time. She presented with 1 week hx of acute psychosis, and was found to be have COVID at the same time. Also treatment-resistant requiring clozapine. No family hx of psychosis. | Y |

| | | | |
|---|---|---|---|
| 34 | Thyrotoxicosis-related psychosis | F/ with known poorly treated thyrotoxicosis. She presented with abnromal behaviour , paranoid idea and hallucination for 10 months. Her FT4 was found to be of very high level upon admission. She was treated with both anitthyroid and antipsychotic medications. Her antipsychotics was stopped after 3 years, and no recurrence of psychosis was known up til the last data entry (3 years after stopping antipsychotics). | N |
| 35 | Uncertain. Ddx: thyrotoxicosis-related psychosis or CNS infection | F/ history of depression, presented with 2 weeks of acute psychosis with fever upon admission. Blood test showed increase free T4 Patient refused lumbar puncture,so she was cinically treated as meningitis based on clinical suspicion. Antithyroid and antipsychotics were also started. Antipsychotics was not stopped after 1 month of remission of psychosis, and she had no recurrence of psychosis up til the point of last data entry (1.5 years after the index episode). | Y |
| 36 | Underlying mitochondrial disease (MELAS) | n/a | N |
| 37 | Organic psychosis (intracranial mass) | F/ hx of anxiety and depression. She presented with 1 day of sudden onset AH and VH, together with 6 months of bilateral decrease in visual acuity. CT brain revealed a large skull base tumor, which eventually confirmed as a pituitary adenoma. Patient refused treatment of her tumor, and refused antipsychotics. | Y |
